# Effects of cognitive-behavioral therapy for insomnia during sedative-hypnotics withdrawal on sleep and cognition in older adults

**DOI:** 10.1101/2025.02.10.25322010

**Authors:** Loïc Barbaux, Nathan E. Cross, Aurore A. Perrault, Mehdi Es-sounni, Caroline Desrosiers, Doris Clerc, Francis Andriamampionona, David Lussier, Cara Tannenbaum, Anik Guimond, Sébastien Grenier, Jean-Philippe Gouin, Thien Thanh Dang-Vu

**Author notes:** corresponding authors;, Department of Health, Kinesiology and Applied Physiology, Concordia University, 7141 Sherbrooke St W, H4B 1R6 Montreal, QC, Canada.

## Abstract

**Objectives:** Our objective was to assess the effect of cognitive-behavioral therapy for insomnia (CBTi) on subjective and objective sleep quality (including sleep spindles) and cognition during a sedative-hypnotics withdrawal program in older adults with insomnia disorder.

**Methods:** We performed a two-arm randomised controlled trial (RCT) of a sedative-hypnotic withdrawal plan alone (WPo group) or combined with CBTi (WP+CBTi group) in 47 older adults with insomnia disorder over a sixteen-week period. Our primary outcomes were change in self-reported insomnia severity (Insomnia Severity Index (ISI)), sleep efficiency (SE) from sleep diaries, and change in SE and spindle density from polysomnographic (PSG) recordings collected at baseline and at post-intervention (16 weeks). Secondary outcomes included other sleep changes from PSG, actigraphy and sleep diaries, sleep and mood questionnaires and neuropsychological assessments (manual dexterity, attention/concentration, verbal inhibition, visuo-spatial abilities).

**Results:** The withdrawal program was effective in achieving discontinuation and reducing insomnia severity, with similar success with and without CBTi. The combined intervention additionally improved subjective sleep quality and prevented the decrease in subjective sleep duration induced by sedative-hypnotic discontinuation. Neither intervention significantly impacted objective sleep architecture or cognitive performance. Furthermore, reduction in sleep spindle density was observed with combined CBTi and withdrawal, but not with withdrawal alone.

**Conclusions:** Both withdrawal alone and sedative-hypnotic withdrawal combined with CBTi effectively facilitated discontinuation and reduced insomnia severity, with the combined intervention further enhancing subjective sleep quality and preserving sleep duration. Although neither approach significantly impacted objective sleep architecture or cognitive performance, the potential reduction in sleep spindle density linked to the combined intervention warrants further investigation.

**STATEMENT OF SIGNIFICANCE:** This study evaluated the combined effects of CBTi and sedative-hypnotic withdrawal on both subjective and objective sleep outcomes, such as sleep spindle density, as well as cognitive performance, in older adults with insomnia disorder. Findings reveal that CBTi, when combined to sedative-hypnotic withdrawal program, not only supports withdrawal success and reduces insomnia severity but also enhances subjective sleep quality and maintains sleep duration, which may be compromised by withdrawal alone. The observed reduction in sleep spindle density, linked to the combined intervention, needs further investigation. These results provide valuable insights into optimizing sedative-hypnotic discontinuation strategies for older adults experiencing chronic insomnia.

## INTRODUCTION

Insomnia disorder, defined by the complaints of difficulties falling and/or maintaining sleep more than 3 nights per week, and/or accompanied by early morning awakenings, despite adequate sleep opportuniges^1^, is highly prevalent in the older population. Insomnia symptoms are the most prevalent sleep disturbance among the elderly, with up to 50% experiencing of difficulty initiating or maintaining sleep^2–5^. Insufficient sleep has been linked to reduced grey matter volume, notably in the thalamus, hippocampus, and cortical areas such as the temporal and orbitofrontal cortices, leading to deficits in attention and working memory^6–9^. It is associated with poorer quality of life and health and increased risks for cognitive decline^10–12^, thereby representing a major health issue.

The chronic consumption of prescribed sedative-hypnotics as a treatment for insomnia is common among the elderly^13–19^. Two major classes of sedative-hypnotics are benzodiazepines (BZD) and benzodiazepine receptor agonist (BZRA)^20^. While BZD/BZRA are effective in reducing subjective insomnia symptoms, many individuals develop tolerance over time^21^. Furthermore, long-term use of these sedative-hypnotics does not result in objective improvements in sleep quality already disrupted by insomnia. The long-term use of these sedative-hypnotics does not lead to objective improvements in the sleep quality already disrupted by insomnia. BZD alter sleep architecture by reducing deep sleep^22,23^ and disrupt the spectral properties of brain oscillations such as spindles and slow oscillations (SO)^24^, which may contribute to impaired memory consolidation and cognitive issues^25^. For instance, both BZD^22,26^ and BZRA^27,28^ use have been shown to increase spindle activity and both sigma and beta power spectrum. In addition, chronic use of BZD^29,30^ and BZRA^31^ are accompanied with dependence and cognitive decline. Furthermore, associations has been found between BZD/BZRA use and the development of dementia^32–34^, but also comorbidities in older adults, including daytime sleepiness and loss of motor coordination, which can increase risk for hip fractures^35^. This is particularly concerning given that approximately 30% of older adults who experienced hip fracture passed away in the subsequent year, while survivors exhibited a gradual decline in quality of life^36^. Considering this evidence, the American Geriatrics Society advises against the use of BZD and BZRA in older adults, regardless of the duration of use^37^.

However, the use of BZD^13–15,17–19^ and BZRA^16^ in older adults remains high, and older adults can become dependent on these medications to be able to sleep, particularly after chronic long-term use. Therefore, it is necessary to encourage withdrawal from BZD/BZRA in older adults to reduce the risk of adverse effects.

Several studies have investigated the impact of sedative-hypnotics withdrawal on sleep quality and cognitive function. BZD/BZRA discontinuation has been shown to improve self-reported sleep and quality of life outcomes in older adults^38^. Moreover, in a sample of 19 adults with insomnia and chronic use of BZD, after a 15-day withdrawal period, they exhibited an improvement in slow wave sleep percentage 15 days after withdrawal^39^. Sleep efficiency declined following withdrawal but gradually improved, returning to levels comparable to pre-withdrawal 15 days post-withdrawal. Furthermore, improvements in attention, concentration, motor performance tasks, verbal and non-verbal memory, and visuo-spatial tasks have been observed after withdrawal in the elderly^40^. However, recovery effects are limited, as residual cognitive deficits remain in most cognitive functions compared to control subjects^40–42^. A major limitation is that BZD/BZRA withdrawal could be unsuccessful and accompanied by a worsening of insomnia symptoms called insomnia rebound^43^. Successful withdrawal is not always easy to achieve in chronic users, and approximately half of them will continue to use sedative-hypnotics after tapering^44,45^. Although withdrawal programs are effective in the short term, the beneficial effects on sleep are not sustained over time, with a gradual return of sleep quality indices to initial values following one year^46^. This is understandable as withdrawal alone is not focused on the fundamental issues associated with the hypnotic consumption, namely chronic insomnia. Therefore, effective withdrawal programs may benefit from implementing other therapeutic approaches to address chronic insomnia.

Cognitive-behavioral therapy for insomnia^47,48^ is considered the first line intervention for the management of insomnia in adults given its long-term efficacy. CBTi is a multimodal psychological intervention aimed at modifying maladaptive thinking and behaviours that contribute to the perpetuation of insomnia^51^ and is highly effective in reducing insomnia severity, improving subjective sleep quality and daytime functioning in diverse populations^52–55^, including older adults^56–58^. Both BZD^44,59,60^ and BZRA^59,60^ withdrawal intervention combined to CBTi program has been shown to reinforce withdrawal success and be more successful in reducing insomnia severity, than standard sedative-hypnotics tapering alone^44,59,60^. However, few studies have assessed the effects of CBTi on both subjective and objective sleep quality during sedative-hypnotic withdrawal in older individuals with chronic insomnia. In addition to investigating subjective sleep quality, describing the impact on objective sleep quality and brain oscillations is important for understanding the implications for cognitive function in older adults.

The objective of this study was to assess the effect of CBTi on sleep quality during a sedative-hypnotics (BZD/BZRA) withdrawal program in older adults with insomnia disorder. Using a RCT design of a sedative-hypnotic withdrawal plan alone (WPo group) or combined with CBTi (WP+CBTi group), our primary outcomes were change in self-reported insomnia severity and sleep quality as well as objective changes in sleep efficiency and spindle density. Secondary outcomes included PSG and sleep diaries-extracted measures of sleep as well as neuropsychological assessments. We expected to observe a greater improvement in sleep objective and subjective outcomes in the combined intervention group (WP+CBTi) than with a sedative-hypnotics withdrawal intervention alone (WPo). We also hypothesized that neuropsychological performances would be greater in the WP+CBTi group compared to the WPo group.

## MATERIAL & METHODS

### Participants

Older adults (≥60 years) chronically using sedative-hypnotics for the management of chronic insomnia were recruited from advertisements (both online and in newspapers), Centre de recherche de l’institut universitaire de gériatrie de Montréal (CRIUGM) participant databases, collaborations with primary care and sleep clinics at IUGM, as well as patient associations. Initially, participants underwent a phone screening to determine their eligibility based on inclusion and exclusion criteria.

Inclusion criteria were: participants aged 60 and older; French speaking; with insomnia disorder; and using sedative-hypnotics (either BZD or BZRA) to treat their insomnia. Participants meeting the DSM-5 diagnostic criteria for insomnia disorder for at least 3 months were included in the study^1^. The DSM-5 criteria for chronic insomnia disorder are defined as self-reported dissatisfaction with sleep associated with initiating sleep (i.e., sleep onset latency greater than 30 min), difficulties maintaining sleep (i.e., wake after sleep onset greater than 30 min), and/ or early morning awakenings (i.e., final awakening time earlier than desired by at least 30 min), for at least 3 times a week and for more than 3 months, combined with significant distress or impairments of daytime functioning, despite adequate sleep opportunities. Participants also had to meet a criterion of sedative-hypnotic use (regardless of the dosage): BZD (Diazepam, Clonazepam, Nitrazepam, Oxazepam, Lorazepam, Temazepam) or BZRA (Zopiclone) drugs had to be prescribed for insomnia and to be used for more than 3 nights a week for more than 3 months. Averaged consumed doses of BZD and BZRA were converted into Diazepam Equivalent Dose, according to the Equivalence Table of BZD in the Ashton Manuel Supplement^61^.

Exclusion criteria were as follows: medical conditions affecting cognition (e.g., Alzheimer or Parkinson’s disease, epilepsy, fibromyalgia, stroke) or sleep (e.g., narcolepsy, sleep apnea with apnea-hypopnea index (AHI) > 15/h, periodic limb movement index (PLMI) during sleep > 15/h, both confirmed by PSG screening); cognitive deficits (dementia or Mini Mental State Examination^62^ – MMSE score ≤ 23); sensorimotor deficits (including visual or auditory deficits); active major depression or psychotic disorders (assessed during a structured clinical interview following the Mini International Neuropsychiatric Interview guidelines^63^); active cancer; night shift work or changes in time zones over the past 2 months; alcohol consumption (> 10 drinks/week) or illicit drug use, currently in palliative care.

### Protocol

Eligible participants were enrolled in a randomised controlled trial comprised of a withdrawal program combined with CBTi-arm (WP+CBTi) and a withdrawal program only control arm ( WPo - see Figure 1A for the study design). Within a month after the initial PSG screening, which also served as an adaptation night, participants completed a baseline assessment (T1). The latter included a second PSG recording, and in the following morning, they completed questionnaires and participated in a neuropsychological assessment. Outside the sleep laboratory, participants completed a two-week sleep diary at home while continuously wearing an actigraphy device throughout this period. Participants were then randomized into two groups using a computer-generated randomisation to ensure balanced groups in terms of age, equivalent dose in Diazepam and the duration of hypnotics use. All participants were then enrolled in the withdrawal program. Participants who underwent CBTi simultaneously with the sedative-hypnotics withdrawal plan constituted the WP+CBTi group (n=26), whereas those who only underwent the withdrawal plan constituted the WPo group (n=21).

**Figure 1:**
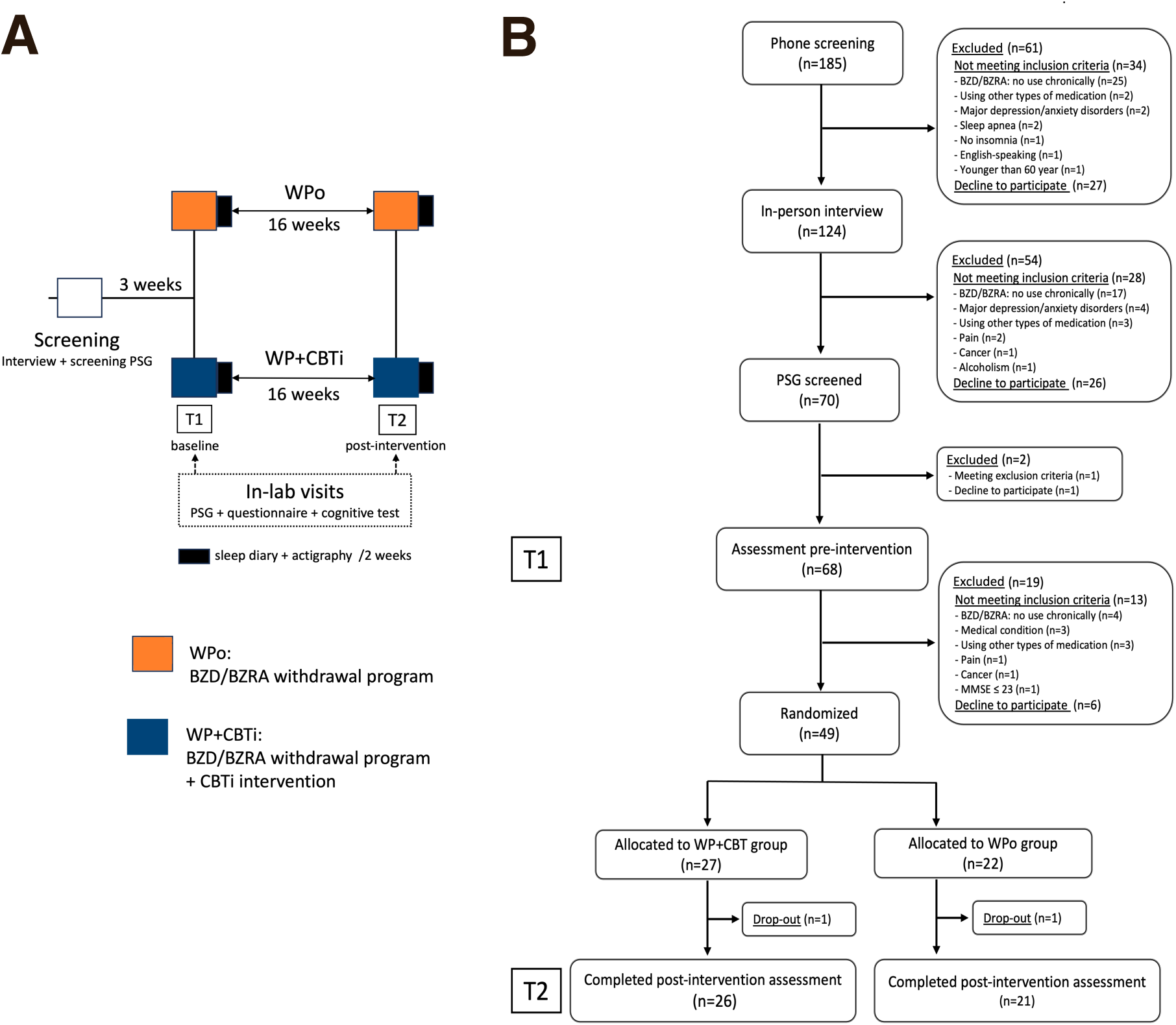
Study Design and participant flow chart. (A) Participant screening included a clinical interview and an overnight PSG to assess exclusion criteria. This first PSG also served as a habituation session to the sleep laboratory environment. At pre-intervention (T1), participants underwent an overnight PSG and the following morning, completed questionnaires and cognitive assessments. Starting from these baseline assessments and continuing over a two-week period, participants completed sleep diaries and wore a wrist actigraphy device. Subsequently, participants were then randomized into either the withdrawal plan combined with CBTi (WP+CBT group) or the withdrawal plan alone group (WPo group). After completing the interventions over a 16-week period (T2), participants underwent the same assessments as during the pre-intervention phase (T1). (B) Participant consort flow chart

Sixteen-week post-randomization (T2), participants from both groups came back to the laboratory and underwent the same protocol as the baseline assessment (PSG, neuropsychological assessment, questionnaires) followed by a 2-week sleep diary and actigraphy (see Figure 1A).

Following post-interventions (T2), participants in the WPo group received the CBTi intervention. All participants signed a written informed consent form, which was approved by the Comité d’Éthique de la Recherche of the CRIUGM. This study was registered as a clinical trial (ISRCTN10037794), https://www.isrctn.com/ISRCTN10037794).

### Withdrawal program

The withdrawal plan included an educational brochure, adapted from and tested in a previous RCT^64^, providing information on the risks of sedative-hypnotic use and tapering, and recommendations for sleep hygiene. This plan was implemented over a sixteen-week period, during which participants received telephone follow-ups every two weeks. These follow-ups aimed to monitor insomnia progression using the ISI^65,66^, assess withdrawal symptoms via the benzodiazepine withdrawal symptom questionnaire^67^, and offer support and encouragement. Reported withdrawal symptoms included physical (e.g., tremors, sweating), emotional (e.g., heightened anxiety, irritability), and cognitive (e.g., concentration difficulties) domains. Symptoms were considered severe if the total score exceeded 20. A visual guide illustrated the gradual reduction of sedative-hypnotic intake, starting with whole tablets, then halving the dosage, followed by quarter-tablets, and alternating doses daily, ultimately leading to complete withdrawal. The achievement of complete or partial withdrawal was self-reported by participants at the post-intervention assessment (T2), where they provided details on the dosage and frequency of sedative-hypnotic use.

### CBTi intervention

The CBTi intervention was delivered to participants in the WP+CBTi group and to those in the WPo group after completing T2 assessments. It was structured over 16-weeks in 8 group sessions of 90-minutes conducted by a trained psychologist. The first four sessions were given every week, and the last four sessions were separated by two weeks apart. The CBTi program consisted of psychoeducation about sleep and circadian rhythms, stimulus control, sleep restriction, sleep hygiene, cognitive therapy, and relaxation based on Morin & Espie^48^. Each group included 3 to 5 participants, for a total of 7 groups. Randomization process employed non-stratified batches to evenly divided the number of participants from WP+CBTi and WPo. If a participant was unable to join one session, a catch-up session was proposed. All participants from the WP+CBTi completed the 8 sessions.

### Measures

#### Questionnaires

Participants completed the following sleep-related questionnaires at pre- and post-intervention (T1 and T2) :

##### Insomnia Severity Index (ISI)

the ISI is a 7 Likert-scale self-assessment questionnaire used to assess the nature, severity, and impact of current insomnia symptoms^65,66^. Each item is scored on a scale of 0 to 4, and the total score varies between 0 and 28 (with higher scores indicating more severe insomnia). It overall Cronbach’s **α** was 0.82. The change in ISI score was one of our primary outcomes for this study. Participants with an ISI score below 8 post-intervention (T2) were classified as remitters, while those with a reduction of 7 or more in their ISI score were considered responders^68^.

##### Pittsburgh Sleep Quality Index (PSQI)

the PSQI is a self-report measure of general sleep quality over the past month. It includes 18 items divided into seven sub-components assessing sleep quality, sleep latency, sleep duration, sleep efficiency, sleep disturbances, medication use, and daytime dysfunction. The global score ranges from 0 to 21, with higher scores indicating poorer sleep quality^69^. It overall Cronbach’s **α** was 0.65.

##### Epworth sleepiness scale (ESS)

the ESS is a self-administered questionnaire designed to measure an individual’s general level of daytime sleepiness during common daily activities. It consists of eight questions, on a 4-point Likert scale. The total score ranges from 0 to 24, with higher scores indicating greater daytime sleepiness^70^. The overall Cronbach’s **α** was 0.71.

##### Geriatric Anxiety Inventory (GAI)

the GAI is a self-report measuring anxiety in older adults^71^. It consists of 20 items, covering various symptoms of anxiety, including worry, tension, and physical symptoms. The total score ranges from 0 to 20, with higher scores indicating more severe anxiety symptoms. The overall Cronbach’s **α** was 0.89.

##### Geriatric Depression Scale (GDS)

the GDS is a self-report measure designed to assess depression in older adults^72^. It consists of 30 items, covering various symptoms of depression, including mood, and cognitive function. The total score ranges from 0 to 30, with higher scores indicating more severe depressive symptoms. The overall Cronbach’s **α** was 0.58.

#### Sleep Diary

At pre- and post-intervention (T1, T2), participants completed the Consensus sleep diary^73^ every morning over two weeks. They reported time spent in bed (TIB), total sleep time (TST), sleep onset latency (SOL), wake duration after sleep onset (WASO), sleep efficiency (TST/TIB*100) and sleep satisfaction (from 1: very bad sleep to 5: very good sleep). For each sleep variable, measures were averaged over 2 weeks at each time point. Only sleep diary data with more than 5 days completed per timepoint were included in the analyses.

#### Actigraphy

At each time point (T1, T2), participants wore a wrist-wearable accelerometer device (Philips Respironics, Murrysville, USA - Actiwatch) for a period of two weeks to extract TIB, TST, SOL, WASO, SE, by averaging the values over the whole period^74,75^. Bedtime and wake-up time estimates were determined visually by identifying periods of no motor activity and light exposure, cross-referenced with sleep schedules recorded in participant’s sleep diaries. For each sleep variable, measures were averaged over 2 weeks at each time point. Only actigraphy data with more than 5 days completed per timepoint were included in the analyses.

#### Polysomnographic (PSG) recording

Whole-night PSG recordings were used at each time point (T1, T2), including electroencephalography (EEG) montage with 15 electrodes (Fz, F3, F4, Cz, C3, C4, Pz, P3, P4, T3, T4, O1, O2, M1, M2) positioned on the scalp according to the 10-20 system AASM guidelines, electromyogram (chin and legs EMG), electrooculogram (EOG), electrocardiogram (ECG). During screening night only, the PSG setup included an oximeter, thoracic and abdominal belts, oral-nasal thermistor, and nasal cannula, to compute the AHI and PLMI and exclude potential sleep disorders. EEG signal was recorded by a Somnomedics amplifier (SomnoMedics, Germany) at a sampling rate of 512 Hz, referenced to Pz online and to contralateral mastoids (M1 and M2) offline for analysis. Only PSG data with > 180min of sleep (TST) recorded per timepoint were included in the analyses.

#### EEG analysis

All sleep scoring was conducted using the Wonambi python toolbox (https://wonambi-python.github.io). EEG analyses were conducted using the Seapipe python toolbox (https://github.com/nathanecross/seapipe). Sleep stages (N1, N2, N3, REM) and wake 30-s epochs scoring as well as the detection of arousal and artefact events were visually scored according to the AASM rules^76^. From the scoring, we computed the calculation at each time point (T1, T2) of: total sleep time (TST; sum of the time spent in different sleep stages), sleep onset latency (SOL; from light off to the first period of sleep), sleep latency to stages (SL), wake after sleep onset (WASO; sum of the nocturnal awakenings duration), time spent in bed (TIB; sum of TST, SOL and WASO), sleep efficiency (SE; TST/TIB*100), arousal density (per epoch of 30 s), sleep fragmentation index (SFI; number of transitions from deep to lighter sleep stages per hour).

Spindles were automatically detected using a validated algorithm^77^ and implemented in the Seapipe toolbox. The spindle detection algorithm consisted of computing the root-mean-square (RMS) of the participant-adapted sigma band with a 0.5 s overlap window and smoothed with Gaussian filter^78,79^. For spindle detection, the highest center frequency peak (integral of the Gaussian fit) over the sigma range specific for each subject was obtained^80,81^. Using those participant-specific adapted sigma ranges, we determined with a 2Hz bandwidth the highest peak in the 10 to 13 Hz range for midline frontal (Fz – slow spindle) and the 13 to 16 Hz range for midline parietal (Pz - fast spindle), as well as the midline central electrode (Cz; 12-15 Hz), on artefact-free derivations accounting for spindle frequency gradient^82,83^. RMS were identified as a spindle event when values exceeded a threshold at 2 SD above the mean peak amplitude. Detection criteria included a spindle duration ranging from 0.5 to 3 s. For spindles detected in NREM (N2 + N3), we extracted the following characteristics: density (i.e., mean number per epoch of 30 s), amplitude (µV), duration (s) and frequency (Hz).

#### Neuropsychological assessment

The neuropsychological assessment focused on cognitive functions usually impaired in chronic BZD users. Six cognitive dimensions were studied based on a previous study investigating partial improvement post-withdrawal^40^. The neuropsychologist was blinded to the participants’ group assignments to avoid bias in evaluating cognitive performance. The neuropsychological assessment lasted approximately one hour and was conducted at pre- and post-intervention (T1, T2), one hour after waking up.

##### Digit Symbol Substitution Test (DSST)

the DSST involves a table with nine numbers, each associated with a geometric symbol^84^. A random series of 140 digits is presented to the participant who must complete it with the corresponding symbols within a maximum of 90 seconds. The number of completed symbols is obtained as a valid measure of attention and concentration, as a scaled score from the Wechsler Adult Intelligence Scale-Third Edition^85^.

##### Rey complex figure & modified Taylor complex figure tests (MTCF/ROCF)

the MTCF/ROCF assess visual-spatial abilities^86,87^. Both tests consist of two phases: a copy phase, where participants draw a complex figure based on a model, and an immediate recall phase, where they reproduce the figure from memory. For each phase, the time taken to complete each drawing is recorded in seconds, and scaled scores and z-scores are obtained. The z-scores are adjusted for sociodemographic variables (age, education level, and sex; z-score SES) and for the copy time, copy score, and immediate recall score (z-score All)^88^. The MCTF and the ROCF are respectively and randomly counterbalanced between each time point (T1, T2).

The Delis-Kaplan executive function system allows the evaluation of cognitive functions in older adults and combined the color/word interference test as the trail making test (TMT), both used to assess executive function^89^.

##### Trail making test (TMT)

the TMT assesses digit and letter recognition alongside visual-motor skills^90^. In the first session (TMT-A), participants connect numbers from 1 to 25, randomly distributed on a sheet, in ascending order as quickly as possible. In the second session (TMT-B), participants alternate between connecting numbers (1 to 13) and letters (A to L) in ascending and alphabetical order, respectively (1-A-2-B, etc.). Completion times in seconds are recorded, and z-scores are calculated, accounting for age and education level^91^.

##### Color/word interference test (Stroop)

this test evaluates attention, verbal inhibition, and flexibility^92^. Initially, participants described the color of cells aloud as quickly as possible (condition 1). Next, they read a series of color names printed in black and white as quickly as possible (condition 2). Then, they were presented with color words printed in a different color (e.g., the word “red” printed in blue) and were asked to identify the color of the ink (condition 3). In the final component, participants stated the word itself when it was framed and the color of the ink when it was not framed (condition 4). For each condition, completion times were recorded in seconds, and scaled scores were obtained from Appendix D of the Examiner Manual.

##### French adaptation of the 16-items free and cued selective reminding test (FCSRT)

the FCSRT is used to measures difficulties in verbal episodic memory^93^. During the encoding phase, sixteen items were presented to the participant who must memorize and retrieved it over immediate and delayed recall, first cued and then free recall. Participant underwent three phases of free and cued recall. The number of words retrieved during the three phases of immediate free and cue recall (with a maximum of 48) and the delayed free and cue recall (with a maximum of 16) were reported. For the free score and the delayed free score recall score, z-score calculated controlling for age, sex and education level^94^.

##### Purdue Pegboard Test (PPT)

the PPT measures manual dexterity and bimanual coordination^95^. Participants are presented with a perforated board containing two rows of twenty-five holes and are instructed to place as many sticks as possible into the holes within thirty seconds, using their dominant hand (condition 1), non-dominant hand (condition 2), and both hands (condition 3). For each condition, the total number of sticks placed is recorded, and z-scores are calculated to account for age and sex^96^.

### Statistical analyses

Statistical analyses were performed using RStudio 1.2.50 (RStudio, Inc., Boston, MA) and R package (ggplot2, sjstats, sjmisc, forcats, emmeans, rstagx, lme4, ltm).

Our primary outcomes were derived from questionnaires (ISI score), 2-week sleep diaries (SE), and PSG-derived measures (SE and spindle density). Secondary outcomes include changes in additional self-reported questionnaire scores, 2-week sleep diary and actigraphy averaged sleep measures, other PSG-derived measures and neuropsychological performances. The sedative-hypnotics withdrawal success was also included, as the percentage decrease of self-reported sedative-hypnotics consumption from baseline (T1) to post-intervention (T2) for each participant, as well as the proportion of participants achieving completion of the withdrawal program in each group.

We used mixed-model analyses of variance (ANOVA), with Time as within-subject factor (T1 vs T2) and Group as between-subject factor (WPo vs WP+CBTi) on every outcome to assess the impact of both interventions. Per-protocol analyses were conducted considering variability in attrition depending on the measures completed. Non-parametric tests were used (WTS, Wald Test Statistic for small sample size) on variables with no homogeneous variance and anormal distribution.

Exploratory bivariate Spearman’s Rho correlations were conducted between the change in sedative-hypnotics dose and changes in primary outcomes in each group to investigate the effects of CBTi in the relationship between change in sedative-hypnotics use and changes in subjective and objective sleep.

Normality of distribution was checked with Shapiro tests and homogeneity of variance was tested with Levene tests. Effect sizes were calculated using Hedges’s g, indicating the degree of change over time (within-group effect) (corrects for small sample size). The level of significance was set to a p-value of <.05 and p-values were adjusted for multiple comparisons (Benjamini-Hochberg/FDR correction). For significant results, both raw (p) and adjusted p-values (q) were reported when necessary.

## RESULTS

Participant demographics are presented in Table 1. Forty-nine individuals with chronic use of sedative-hypnotics for chronic insomnia were randomized to either the withdrawal program combined to CBTi (WP+CBTi group – N= 27) or the withdrawal program only (WPo group – N= 22). Participants were older adults (69.1 ± 6.2 y.o) chronically using BZD/BZRA (dose equivalent in Diazepam: 7.1 ± 7.9 mg/week; duration: 9.7 ± 8.0 years). They were in majority female (70.2%) and most held a university diploma (mean 14.7 ± 2.4 education years). The majority of participants used BZRA (Zopiclon: 68%), while a smaller proportion used BZD, including Oxazepam: 13%, Lorazepam: 9%, Clonazepam: 4%, Nitrazepam: 4% and Temazepam: 2%.

**Table 1:** Demographics

| Characteristics | WP+CBTi group |  | WPo group |  |
| --- | --- | --- | --- | --- |
| | range | Mean ± SD | range | Mean ± SD |
| N |  | 26 |  | 21 |
| Age | [61-90] | 70.39 ± 6.4 | [61-84] | 67.52 ± 5.8 |
| Sex |  |  |  |  |
| Female \| Male |  | 18 \| 8 |  | 15 \| 6 |
| % | 69.2 | 30.8 | 71.4 | 28.8 |
| Education years | [8 - 19] | 14.5 ± 2.5 | [11 - 20] | 14.9 ± 2.3 |
| Sedative-hypnotic consumption |  |  |  |  |
| Duration [years] | [1 - 23] | 9.6 ± 7.1 | [1 - 35] | 9.8 ± 9.2 |
| Dose equivalent in Diazepam [mg/week] | [0.8 - 25] | 6.4 ± 5.7 | [2.5 - 59] | 8.0 ± 10.3 |
| Score at T1 |  |  |  |  |
| Psychiatric comorbidities |  |  |  |  |
| ESS [/24] | [0 - 18] | 4.8 ± 3.9 | [2 - 10] | 5.0 ± 2.7 |
| GAI [/20] | [0 - 17] | 7.4 ± 5.3 | [0 - 17] | 6.2 ± 5.2 |
| GDS [/30] | [0 - 18] | 7.4 ± 5.1 | [1 - 21] | 9.3 ± 5.8 |
| MMSE [/30] | [24 - 30] | 27.8 ± 1.9 | [25 - 30] | 28.3 ± 1.7 |
| ISI [/28] | [2 - 23] | 12.5 ± 5.0 | [7 - 26] | 15.2 ± 5.0 |
| PSQI [/21] | [4 - 18] | 10.7 ± 3.7 | [7 - 18] | 11.4 ± 3.3 |

ESS, Epworth Sleepiness Scale; ISI, Insomnia Severity Index; GAI, Geriatric Anxiety Inventory; GDS, Geriatric Depression Scale; MMSE, Mini Mental State Examination; PSQI, Pittsburgh Sleep Quality Index; WPo, sedative-hypnotics withdrawal plan group; WP+CBTi, CBTi combined to sedative-hypnotics withdrawal plan

We did not find any difference between the WP+CBTi and WPo groups regarding age, sex, education years, sedative-hypnotics dose and duration, as well as anxiety, depression and sleep quality (ISI, PSQI) levels (t-test, Wilcoxon test, Fisher test; all *p*>.05).

### Participant adherence and primary outcome attrition

During the intervention, 2 participants (N=1 from WP+CBTi; N=1 from WPo) dropped out of their participation to the project (see Figure 1B). At 16 weeks post-randomization (T2), 26 participant (attrition rate 3.7%) from the WP+CBTi group and 21 participants (attrition rate 4.6%) from the WPo group filled the ISI questionnaire (primary outcome). Regarding self-reported SE, 25 participants (attrition rate 7.4%) from the WP+CBTi group and 19 participants (attrition rate 9.5%) from the WPo group filled sleep diaries at 16 weeks post-randomization (T2). Regarding objective SE and spindle density obtained from PSG recordings, 25 participants (attrition rate 7.4%) from the WP+CBTi group and 20 participants (attrition rate 9.1%) from the WPo group completed overnights at 16 weeks post-randomization (T2). Attrition rates of the secondary measures can be found in Supplemental materials.

#### Effect of both interventions on self-reported sedative-hypnotic discontinuation

A Time effect was found on the sedative-hypnotic dose (*F*(2,47)=135, *p*<.001), where the self-reported dose was significantly reduced in both the WPo (dose T2 - T1: −4.6 ± 3.4 mg/week; *p*<.001) and the WP+CBTi groups (dose T2-T1: −5.6 ± 5.8 mg/week; *p*<.001) (see Figure 2A and Table 2). We did not find significant differences between groups following sedative-hypnotic discontinuation (T2); and no interaction between Time and Group (all *p*>.05). We also did not find group differences in the percentage of reduction in sedative-hypnotic consumption (WPo: 82.8 ± 32.2 %; WP+CBTi: 81.7 ± 30.6 %; *p*=.73), or in the proportion of participants achieving complete tapering (WPo: 71.4 ± 4.6 %; WP+CBTi: 65.4 ± 4.9 %; *p*=.76).

**Figure 2:**
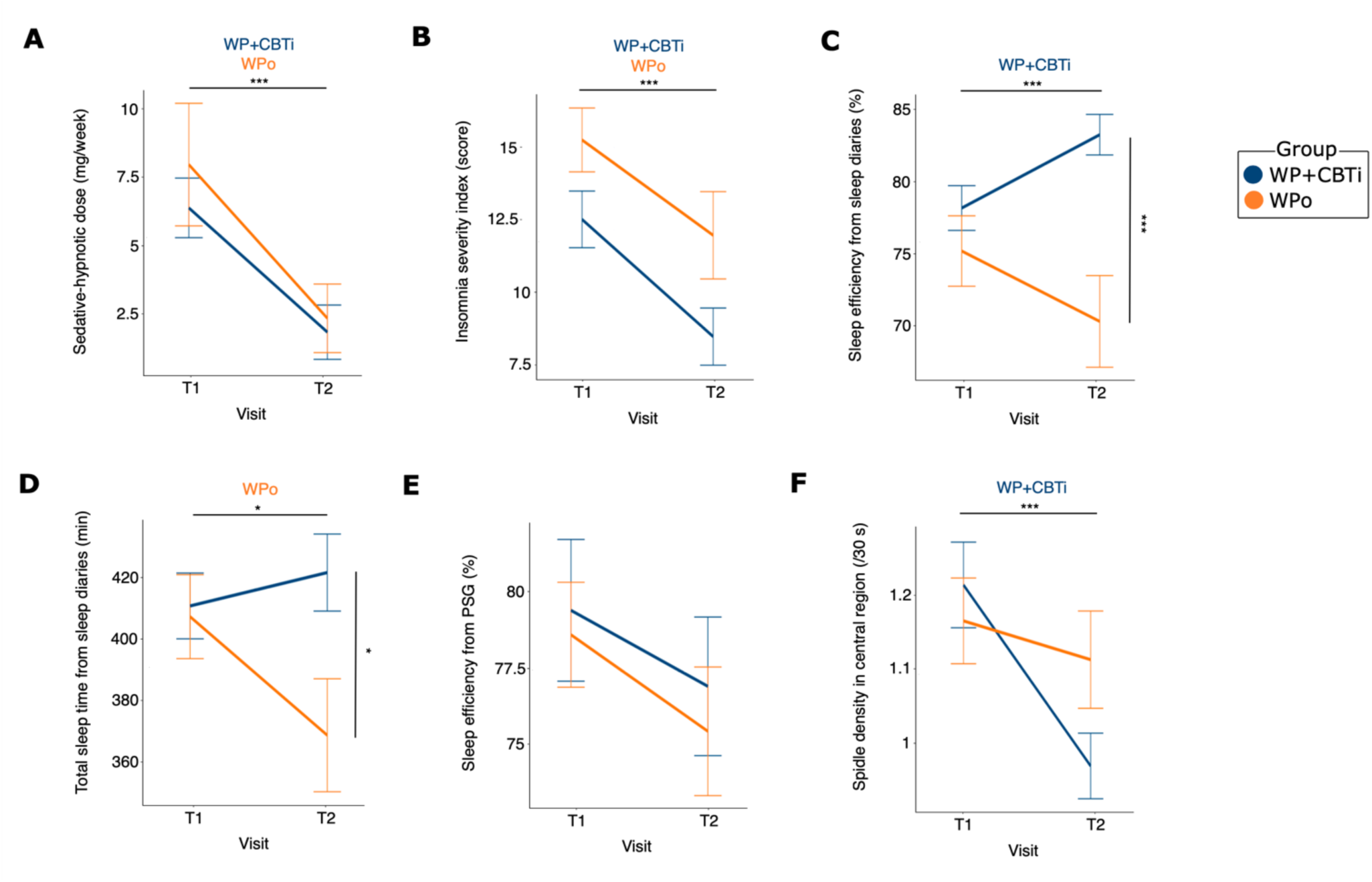
Sedative-hypnotic dose and sleep parameters changes at post-intervention. (A) Sedative-hypnotic dose consumption was reduced post-intervention for both groups (B) Insomnia severity was reduced post-intervention in both groups (C) Sleep efficiency from sleep diaries increased in CBTi with difference between groups at T2 following withdrawal (D) Total sleep time from sleep diaries decreased in withdrawal alone with difference between groups at T2 (E) Sleep efficiency from polysomnography (PSG) did not change post-intervention (F) Spindle density in central region was reduced in CBTi following withdrawal Asterisks represent significance (p): *<.05; **<.01; ***<.001

**Table 2:**
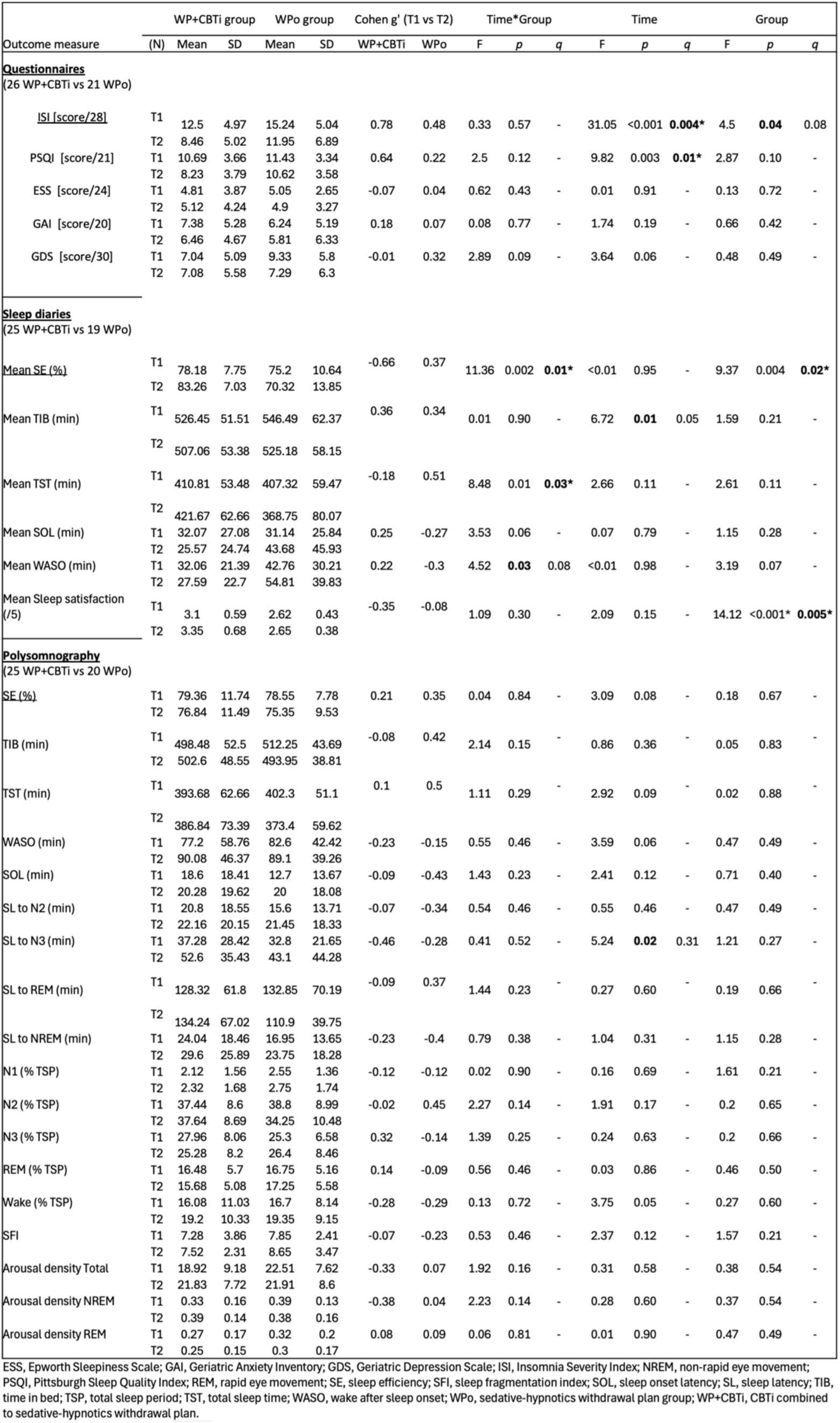
Self-reported sleep quality from diaries, questionnaires and PSG-related outcomes

In the WP+CBTi group, out of 27 participants, 1 (3.7%) was not successful at reducing their consumption of sedative-hypnotics, while 2 participants (9%) from the WPo group did not reduce their BZD/BZRA use. Additional analyses indicate that sedative-hypnotic dose at baseline is negatively associated with the percentage reduction in self-reported consumption after both interventions (WP+CBTi: *r*=-.5; *p*<.001 - Figure S1; WPo: *r*=-.3; *p*=.04). Furthermore, higher levels of depression (WP+CBTi: *r*=.01; *p*=.97; WPo: *r*=-.5; *p*=.03) and anxiety (WP+CBTi: *r*=-.07; *p*=.75; WPo: *r*=-.6; *p*=.002) at baseline (T1) were linked to a lower success rate in withdrawal in the WPo group only.

#### Effect of both interventions on subjective sleep quality

At 16-week post-randomization, per-protocol analysis showed a Time effect for ISI score (*F*(2,47)=31.1, *p*<.001, *q*=.004 – Figure 2B), where both the WP+CBTi and the WPo groups displayed reduced insomnia severity at T2 compared to T1 (all *p*<.001) (see Table 2). The WPo group exhibited a moderate change in ISI scores (g’=.5), whereas the WP+CBTi group showed a large change (g’=.8). A Group effect (*F*(2,47)=4.5, *p*=.04, *q*=.08) was found, however, it did not survive correction for multiple comparisons. No significant interaction between Time and Group was observed (*p*>.05). Post-hoc analyses did not show significant Group difference at T1 and T2 (all *p*<.05). In the WP+CBTi group, 7 out of 26 participants (26.9%) were considered as responders (ISI score reduced by at least 8 point at T2) and 12 (46.2%) were considered in remission (ISI score below 8 at T2). Meanwhile, in the WPo group, 3 out of 21 participants (14.3%) were considered as responders and 7 (33.3%) were considered in remission. There were no significant Group differences in the number of participants responders or in remission at T2 (all *p*>.05).

Concerning change in PSQI score (WP+CBTi N=26; WPo N=21), we found a Time effect (*F*(2,47)=9.8, *p*=.003, *q*=.01– Figure 4) driven by a significant reduction in PSQI score in WP+CBTi group only (−23%, *p*=.003, g’=.6; WPo: −9% *p*>.05, g’=.2). However, we found no Time*Group interaction (*F*(2,47)=2.5, *p*>.05) or Group effect (*F*(2,47)=2.9, *p*>.05).

No associations were observed between change in sedative-hypnotic dosage and change in ISI or PSQI scores in both groups (all *p*>.05).

Regarding self-reported SE from sleep diary (WP+CBTi N=25; WPo N=19), per-protocol analysis revealed a significant Group*Time interaction (*F*(2,44)=11.4, *p*=.002, *q*=.008) as well as a Group effect (*F*(2,44)=9.4, *p*=.004, *q*=.02) (see Figure 2C - Table 2). The increase in SE was only observed in the WP+CBTi group (+6.5%; *p*=.004, g’=-.7; WPo: −6.5%, *p*>.05, g’=.4) with a significant difference between groups at T2 (*p*=.001). There was a significant Group*Time interaction (*F*(2,44)=8.5, *p*=.006, *q*=.03) but no Group or Time effect (all *p*>.05) for self-reported sleep duration due to a decrease in TST in the WPo group only (−9.5%; *p*=.02, g’=.5; WP+CBTi: +2.6%, *p*>.05, g’=-.2) with a significant difference between the groups at T2 (*p*=.02) (see Figure 2D). While we found no Time*Group interaction (*F*(2,44)=1.1, *p*>.05) or Time effect (*F*(2,44)=2.1, *p*>.05), we observed a significant Group effect (*F*(2,44)=14.1, *p*<.001, *q*=.005) on sleep satisfaction, where post-hoc tests indicated significant differences between groups at both T1 (*p*=.01) and T2 (*p*=.001). We found a significant Group*Time interaction on WASO, although it did not pass multiple comparison (*F*(2,44)=4.5, *p*=.03, *q*=.08), due to an increase WPo group only (−28.2%; *p*=.04, g’=-.3; WP+CBTi: −13.9%, *p*>.05, g’=.2) with a significant difference between groups at T2 (*p*=.02). Analyses of self-reported questionnaire scores (GAI, GDS, ESS) and other measures of sleep (e.g., wake duration, latency) using sleep diaries did not reveal any significant interaction or main effect of Time or Group (all *p*>.05) and displayed none-to-small effect size (g’<0.3; see Table 2). No association was found between change in SE and change in sedative-hypnotic dosage at post-intervention in both groups (all *p*>.05).

#### Effect of both interventions on objective sleep

Per-protocol analysis from PSG-related measures (WP+CBTi N=15; WPo N=12) revealed no changes in the primary outcome SE (*p*<.05) (see Figure 2E). We also did not find any effects of Group and Time on additional sleep architecture measures or averaged sleep measure extracted from actigraphy (WP+CBTi group: n=25; WPo group: n=20) (all *p*<.05) (see Tables 2 and S1). Further analyses per sleep stage N2 and N3 separately, are also presented in Supplemental Material.

#### Effect of both interventions on sleep spindle characteristics

Regarding changes in sleep spindle density (primary outcome), per-protocol analysis (WP+CBTi N=23; WPo N=20) indicated a significant Time effect of spindles detected during NREM sleep in the central region (F(2,43)=8.9, *p*=.005, *q*=.01; see Figure 2F), primarily driven by a reduction in spindle density in the WP+CBTi group only (−19.8%, *p*<.004, g’=1; WPo: −5.1%, *p*>.05, g’=.2) (see Table S3). Similar results were found per stage (see Supplemental Material).

Further analyses revealed that the decrease in central spindle density in WPo correlated with the reduction in the sedative-hypnotic dose consumed (*r*=.5; *p*=.04 - Figure S2). However, similar correlation was not found in the WP+CBTi group (*p*>.05).

We observed no interaction or main effects of Group or Time on central spindle characteristics (i.e., duration, amplitude, peak frequency).

### Effect of both interventions on cognitive performance

There were no significant interactions or main effects of Time or Group with none-to-small effect size on neuropsychological performance (all *p*<.05 and g’<.2) (see Table S2).

## DISCUSSION

This two-arm RCT investigated the effects of CBTi on both subjective and objective measures of sleep quality, as well as cognitive function, during sedative-hypnotic withdrawal in older individuals with chronic insomnia. Both groups resulted in a significant reduction in sedative-hypnotic dosage, with participants achieving an 80% decrease post-intervention. Both groups also decreased insomnia severity from baseline to follow-up. Additionally, the combination of CBTi intervention and sedative-hypnotic withdrawal enhanced self-reported sleep quality (SE) from sleep diaries and prevented a decline in sleep duration (TST) and the trend toward increased nocturnal awakenings (WASO) observed after withdrawal alone. Neither intervention resulted in changes to cognition or sleep architecture objective assessment. However, both interventions were associated with a reduction in spindle density. The withdrawal program successfully achieved sedative-hypnotic withdrawal. Similarly, previous research showed that older adults with insomnia experienced equivalent success rates when discontinuing sedative-hypnotics, whether through withdrawal alone or combined with CBTi^97,98^ or self-help CBTi^99^ at post-intervention. Our findings demonstrate that gradual dose reduction is effective in reducing sedative-hypnotic use, with CBTi not appearing to enhance an already high withdrawal success rate. The high success rate in the withdrawal alone condition suggests a potential ceiling effect, limiting the impact of CBTi on further improvement^97–99^.

The withdrawal intervention, without the inclusion of CBTi, likely benefited from regular telephone follow-ups, which provided support, reinforced motivation and contributed to the success of the withdrawal process. This could be due to the fact that CBTi is not primarily designed to target sedative-hypnotic consumption^91,92^. Other factors also contribute to withdrawal success. For instance, a lower dose of sedative-hypnotics at baseline was associated with a higher discontinuation rate post-intervention, which is consistent with previous findings^101^. In the present study, participants with high baseline anxiety and depression showed the lowest withdrawal success in withdrawal alone. Psychological distress, readiness to change, and self-efficacy also influence sedative-hypnotic discontinuation in individuals with chronic insomnia^102^. However, motivation may still play a role in the challenges associated with withdrawal success^103^.

We observed a reduced insomnia severity following sedative-hypnotic withdrawal, with a moderate effect for withdrawal alone and a large effect when combined with CBTi. In line with these findings, a prior study reported a similar reduction in ISI scores among older adults who received CBTi, either alone or combined with sedative-hypnotic tapering, compared to withdrawal alone and sustained at the 12-month follow-up^96^. Other studies did measure insomnia severity and observed a reduction following sedative-hypnotic withdrawal, either alone or in combination with CBTi^97^. These findings challenge the long-term efficacy of BZD and BZRA in managing insomnia, as their discontinuation did not overall exacerbate symptoms in our participants. Instead, insomnia severity declined. Likewise, sedative-hypnotics withdrawal had no adverse impact on anxiety^104^. While in this study, insomnia severity improved post-withdrawal regardless of CBTi therapy, there was a stronger benefit after CBTi. The positive impact of combining CBTi and withdrawal on insomnia severity may become more apparent over time^104^. For example, in older adults receiving sedative-hypnotic withdrawal alone, the occurrence of rebound of insomnia symptoms was found at post-intervention; however, self-reported sleep quality improved at the follow-up six-months later^38^. However, our study design did not include a long-term follow-up assessment to determine whether CBTi could be more effective in managing insomnia and preventing relapse. Future studies should further describe the long-term effect of combined interventions on self-reported sleep quality and insomnia severity.

Nevertheless, the combination of CBTi and withdrawal showed benefits by improving self-reported sleep quality, as seen in the reduced PSQI score and increased SE from sleep diaries. No such improvements were observed with withdrawal alone. Another study found that both the combined intervention and CBTi intervention alone leaded to an increased SE from sleep diaries in older adults^32^. The beneficial effect of combined interventions on self-reported sleep quality was also reported in adult and middle-aged adults with chronic insomnia through questionnaire or sleep diaries^38^. Furthermore, the long-term benefits of CBTi were sustained at one- and two-year follow-ups, whether delivered independently or alongside sedative-hypnotic tapering, in older adults with chronic insomnia^106^. Yet, it has also been observed that sedative-hypnotic discontinuation in adults with chronic insomnia also leads to self-reported improvements in sleep quality, as recorded in sleep diaries two-weeks post-intervention^39^. However, sleep quality scores remained lower compared to those reported by controls without sleep disorders. Overall, the evidence suggests that CBTi interventions may improve various subjective sleep dimensions following withdrawal of sedative-hypnotics, enhancing both overall sleep quality (PSQI) and insomnia severity (ISI).

In addition, CBTi effectively prevented the decline in sleep duration typically observed with sedative-hypnotic withdrawal. However, the significant change in TST was observed only in withdrawal alone, despite no change in SE. Notably, sedative-hypnotic withdrawal, whether combined with CBTi or as a standalone intervention, has been reported to reduce TST while improving SE^44^. By including strategies to consolidate sleep by aligning TIB more closely with actual sleep needs, CBTi improves the quality of sleep rather than its duration, which explains why SE improves without a corresponding increase in TST. In contrast, older adults undergoing tapering, either alone or with CBTi, showed increased TST and SE^97^, highlighting the need for further research on how combining CBTi with withdrawal affects sleep duration.

The withdrawal program combined with CBTi did not improve objective sleep quality, as measured by PSG recordings and actigraphy data post-intervention, nor did either intervention alone. Aligning with the general CBTi research, this finding is not surprising. CBTi interventions did not have an effect on objective PSG measures in middle-aged adults with chronic insomnia^53^. Meta-analyses suggest that improvements in sleep quality following CBTi are more consistently detected through self-reported assessments than through objective measures like actigraphy or PSG^50,107^.

Objective assessment of sleep quality via PSG after a CBTi intervention are limited in the literature. Only small changes were found in PSG outcomes following sedative-hypnotic withdrawal, whether alone or combined with placebo biofeedback or CBTi. Specifically, reduced TST and SE were observed following all three interventions^97^. Another study involving older adults receiving a withdrawal plan with or without CBTi reported increased N3 and REM duration, while TST and N2 duration decreased post-intervention, although no significant group effect was observed^44^. Similar to subjective sleep, is possible that improvements in objective sleep quality may emerge with a delay, as one study showed that sleep quality could improve two weeks after sedative-hypnotic withdrawal^39^. However, these improvements were not sustained over time, as sleep quality returned to baseline after one-year follow-up^44,97^.

The CBTi intervention combined to sedative-hypnotic withdrawal was effective in reducing spindle density in the central region. Chronic BZD use has been found to increase spindle density in adults^26^, which may explain the observed decrease in spindle density following discontinuation of long-term sedative-hypnotic use. Although such a reduction was not observed following sedative-hypnotic withdrawal alone, the decrease in spindle density correlated with the reduction in the sedative-hypnotic dose consumed. This is the first study to report this effect, the clinical significance of which remains to be further investigated.

No effects on cognitive functioning were observed following sedative-hypnotics withdrawal combined to CBTi and withdrawal alone. In the short-term, CBTi has been shown to not change objective cognitive assessment^108^. In middle-aged adults with chronic insomnia, CBTi did not influence either the objective or subjective measures of cognitive functioning^53^. A study reported an increase in self-reported cognitive function following CBTi in middle-aged adults, while no improvement was captured through objective cognitive assessments^109^. Discrepancies exist between subjective reports of daytime impairment — commonly reported among individuals with chronic insomnia — and objective neuropsychological performance^110^. This may account for the lack of cognitive changes following both interventions. There is a lack of research investigating the ideal post-intervention assessment period that is necessary to capture cognitive improvements that may emerge over a longer timeframe. Literature is also lacking in describing the effects of combining CBTi with sedative-hypnotics withdrawal, as well as withdrawal alone, on cognition. Sedative-hypnotic withdrawal in older adults with chronic use improved information processing speed and accuracy at post-intervention^41^. A meta-analysis reported cognitive improvements following sedative-hypnotic withdrawal in chronic users^40,42^. However, impairments, particularly in verbal memory persisted when compared to controls or normative data. Furthermore, sedative-hypnotics withdrawal in older adults was found associated with prolonged impairment in attentional and psychomotor cognitive functions, sustained for at least six months post-intervention^45^.

Following the withdrawal intervention, both groups showed equivalent outcomes, regardless of CBTi inclusion. While the response rate (>7-point reduction in the ISI) was low (26%), remission (ISI score <8 at post-intervention) was high (46%). This suggests that remission occurred even without reaching the typical score reduction threshold used to define treatment response. This likely reflects that many participants did not have severe insomnia at baseline (ISI < 22).

The findings may be influenced by several limitations. First, the sample size was limited and consisted of a majority of females (F = 70%), not allowing for sex differences analysis. Second, the description of the combined intervention’s impact on outcomes may require adding a CBTi intervention alone group, which was not included in this study. Third, the dosage of sedative-hypnotics was self-reported, and we did not conduct urine toxicology screening to objectively assess the dose used and could lead to inaccurate withdrawal success rate. Fourth, missing data limited complete observation. For example, discrepancies were noted between the changes in SE from self-report SE and actigraphy, limited analysis of sleep misperception—which has been shown to be significantly impact by CBTi^53^. Finally, the long-term effects of the combined intervention on sleep quality and cognition were not assessed here.

In conclusion, effective sedative-hypnotic withdrawal resulted in a reduction in insomnia severity. When combined with a CBTi intervention, this additionally improved subjective sleep quality and prevented decreases in sleep duration induced by sedative-hypnotic discontinuation. Neither intervention, however, significantly impacted objective sleep architecture or cognitive performance. Furthermore, reductions in sleep spindle density may be attributed to CBTi, but further investigation is needed to clarify these findings.

## Data Availability

All data produced in the present study are available upon reasonable request to the authors

## AUTHOR CONTRIBUTION STATEMENT

Conceptualization: LB, NEC, TDV, AAP

Recruitment: ME, LB, DC, CD, FA, DL, SG, JPG, CT

Data collection: LB, ME

Project coordination: LB, ME

Data curation: LB, ME, NEC

Formal analysis: LB

Methodology: JOB, NEC, AAP

Visualization: LB, NEC, AAP

Interpretation: LB, NEC, AAP, TDV

Supervision: AAP, NEC, TDV

Funding: TDV, JPG

Writing - original draft: LB, NEC, AAP, TDV

Editing and reviewing: JPG, CT, SG, DL, CD, FA, ME, DC

## ACKNOWLEDGMENTS

This research was supported by grants from the Canadian Institutes of Health Research (MOP 142191, PJT 153115) to TDV and JPG, from the Natural Sciences and Engineering Research Council of Canada to TDV, from the Centre de Recherche de l’Institut Universitaire de Gériatrie de Montréal and from Concordia University to TDV. SG and TDV are supported by a Senior Research Scholar award from the Fonds de recherche du Québec-Santé (FRQS). LB has been supported by the CIHR-SPOR Chair in Innovative, Patient-Oriented, Behavioural Clinical Trials, Concordia University.

We acknowledge the contributions of the following team members who assisted in participants’ recruitment, data collection and data preprocessing: students Jean-Louis Zhao, Romain Perera, Ali Salimi, Cristina Bata, Emma-Maria Phillips, and Julia Giraud, postdoctoral fellow Mathilde Reyt; research associate Florence B. Pomares; and all the volunteers. We also thank our sleep technologists Madeline Dickson and Elinah Mozhentiy from the Clinique SomnoMed for their contribution to the setup of sleep recordings, and Suzanne Gilbert for her contribution to the study conceptualization. Finally, we would like to thank the participants for giving their time and energy into this research study.

## ABBREVIATIONS

AASM: American Academy of Sleep Medicine
AHI: Apnea-hypopnea index
BZD: Benzodiazepine
BZRA: Benzodiazepine Receptor Agonist or Z-drugs
CBTi: Cognitive-behavioral therapy for insomnia
CRIUGM: Centre de recherche de l’institut universitaire de gériatrie de Montréal
DSST: Digit Symbol Substitution Test
DKEFS: Delis-Kaplan executive function system (Stroop)
ECG: Electrocardiogram
EEG: Electroencephalogram
EMG: Electromyogram
EOG: Electrooculogram
ESS: Epworth sleepiness scale
FCSRT: French adaptation of the 16-items free and cued selective reminding test
GABA: Gamma-aminobutyric acid
GAI: Geriatric anxiety inventory
GDS: Geriatric depression scale
Hz: Hertz
ICSD: International Classification of Sleep Disorder
ISI: Insomnia severity index
MINI: Mini-international neuropsychiatric interview
MMSE: Mini-mental state examination
MoCA: Montreal cognitive assessment
MTCF/ROCF: Rey complex figure & modified Taylor complex figure tests
NREM: Non-rapid-eye-movement
N1: NREM stage 1
N2: NREM stage 2
N3: NREM stage 3
PLMI: Periodic limb movement index
PPT: Purdue Pegboard Test
PSG: Polysomnography
PSQI: Pittsburgh sleep quality index
REM: Rapid-eye-movement
RCT: Randomised controlled trial
SD: Standard deviation
SE: Sleep efficiency
s: Second
SES: sociodemographic variables (age, education level, and sex)
SFI: Sleep fragmentation index
SO: Slow oscillation
SL: Sleep latency
SOL: Sleep onset latency
TIB: Time spent in bed
TMT: Trail Making Test
TST: Total sleep time
WASO: Wake after sleep onset
WP+CBTi: Withdrawal Plan + CBTi Group
WPo: Withdrawal Plan alone Group
μV: microvolts

## Notes

### Competing Interest Statement

The authors have declared no competing interest.

### Clinical Trial

ISRCTN10037794

### Author Declarations

the Comite d Ethique de la Recherche of the Institut Universitaire de Geriatrie de Montreal gave ethical approval for this work

