## Supplementary materials for "Effects of cognitive-behavioral therapy for insomnia during sedative-hypnotics withdrawal on sleep and cognition in older adults"

Barbaux et al.

### Tables

#### Table S1: Actigraphy-derived outcomes

| Outcome measure | (N) | WP+CBT group Mean | SD | WPo group Mean | SD | Cohen g' (V1 vs V2) WP+CBT | Wpo | Time*Group F | *p* | *q* | Time F | *p* | *q* | Group F | *p* | *q* |
|---|---|---|---|---|---|---|---|---|---|---|---|---|---|---|---|---|
| **Actigraphy** (15 WP+CBT vs 12 WPo) | | | | | | | | | | | | | | | | |
| Mean TIB (min) | V1 | 516.54 | 40.57 | 513.63 | 51.99 | 0.08 | -0.01 | 0.71 | 0.40 | - | 0.14 | 0.71 | - | 0.01 | 0.91 | |
| | V2 | 510.27 | 74.03 | 514.03 | 41.09 | | | | | | | | | | | |
| Mean TST (min) | V1 | 446.89 | 42.7 | 446.74 | 49.48 | 0.38 | 0.35 | 0.19 | 0.66 | - | 4.8 | **0.03** | 0.15 | 0.01 | 0.93 | - |
| | V2 | 414.75 | 92.06 | 413.82 | 98.16 | | | | | | | | | | | |
| Mean SOL (min) | V1 | 15.95 | 18.85 | 13.18 | 11.35 | 0.01 | -0.21 | 0.12 | 0.73 | - | 1.29 | 0.26 | - | <0.01 | 0.98 | - |
| | V2 | 15.81 | 11.2 | 17.71 | 19.68 | | | | | | | | | | | |
| Mean WASO (min) | V1 | 48.92 | 20.3 | 44.22 | 16.26 | 0.37 | -0.06 | 0.99 | 0.32 | - | 0.54 | 0.47 | - | <0.01 | 0.97 | - |
| | V2 | 41.18 | 19.34 | 45.4 | 20.66 | | | | | | | | | | | |
| Mean SE (%) | V1 | 86.01 | 5.98 | 87.08 | 4.44 | 0.38 | 0.32 | 0 | 0.95 | - | 2.79 | 0.10 | - | 0.01 | 0.92 | - |
| | V2 | 82.17 | 11.97 | 79.97 | 18.27 | | | | | | | | | | | |

TIB, time in bed; TST, total sleep time; SOL, sleep onset latency; WASO, wake after sleep onset; SE, sleep efficiency; WPo, sedative-hypnotics withdrawal plan group; WP+CBT, CBTi combined to sedative-hypnotics withdrawal plan

Regarding actigraphy measures, at 16 weeks post-randomization (T2), 15 participants (attrition rate 44.4%) from the WP+CBTi group and 12 participants (attrition rate 45.5%) from the WPo group.

#### Table S2: Cognitive assessment performance

| Outcome measure | (N) | WP+CBTi group Mean | SD | WPo group Mean | SD | Cohen g' (T1 vs T2) WP+CBT | WPo | Time*Group F | *p* | *q* | Time F | *p* | *q* | Group F | *p* | *q* |
|---|---|---|---|---|---|---|---|---|---|---|---|---|---|---|---|---|
| **Cognitive functioning** (25 WP+CBTi vs 21 Wpo) | | | | | | | | | | | | | | | | |
| *Manual dexterity* | | | | | | | | | | | | | | | | |
| PPT - Condition 1 (z-score) | T1 | -0.11 | 1.11 | -0.11 | 1.07 | 0.15 | -0.4 | 1.04 | 0.31 | - | 0.83 | 0.36 | - | 0.82 | 0.37 | - |
| | T2 | -0.3 | 1.34 | 0.33 | 1.07 | | | | | | | | | | | |
| PPT - Condition 2 (z-score) | T1 | -0.12 | 0.99 | -0.17 | 0.71 | -0.23 | -0.65 | 1.25 | 0.27 | - | 7.53 | **0.01** | 0.16 | 0.24 | 0.63 | - |
| | T2 | 0.12 | 1.03 | 0.4 | 0.95 | | | | | | | | | | | |
| PPT - Condition 3 (z-score) | T1 | 0.21 | 1.36 | 0.15 | 0.93 | 0.01 | -0.43 | 0.03 | 0.87 | - | 2.43 | 0.12 | - | 0.51 | 0.47 | - |
| | T2 | 0.19 | 1.27 | 0.61 | 1.13 | | | | | | | | | | | |
| *Attention/concentration* | | | | | | | | | | | | | | | | |
| DSST (scaled score) | T1 | 8.36 | 2.12 | 8.48 | 1.91 | <0.01 | -0.36 | 2.28 | 0.13 | - | 3.78 | 0.05 | - | 2.47 | 0.12 | - |
| | T2 | 8.36 | 1.93 | 9.19 | 1.86 | | | | | | | | | | | |
| *Visual-motor skills* | | | | | | | | | | | | | | | | |
| TMT-A (z-score) | T1 | 0.36 | 1.05 | 0.33 | 1.42 | 0.16 | 0.15 | 0.11 | 0.74 | - | 0.03 | 0.87 | - | 0.42 | 0.51 | - |
| | T2 | 0.19 | 0.89 | 0.13 | 1.01 | | | | | | | | | | | |
| TMT-B (z-score) | T1 | 0.86 | 1.43 | 0.87 | 2.25 | 0.01 | 0.15 | 0 | 0.99 | - | 0.11 | 0.74 | - | 0.6 | 0.44 | - |
| | T2 | 0.83 | 1.65 | 0.57 | 1.63 | | | | | | | | | | | |
| *Verbal inhibition and flexibility* | | | | | | | | | | | | | | | | |
| DKEFS - Condition 1 (scaled score) | T1 | 10.44 | 3.08 | 11.33 | 1.91 | -0.01 | -0.25 | 1.75 | 0.19 | - | 3.64 | 0.06 | - | 1.46 | 0.23 | - |
| | T2 | 10.48 | 3.24 | 11.86 | 2.13 | | | | | | | | | | | |
| DKEFS - Condition 2 (scaled score) | T1 | 10.76 | 2.44 | 11.9 | 1.84 | 0.03 | 0.03 | 0.03 | 0.85 | - | 0.08 | 0.78 | - | 3.51 | 0.06 | - |
| | T2 | 10.68 | 2.41 | 11.86 | 1.68 | | | | | | | | | | | |
| DKEFS - Condition 3 (scaled score) | T1 | 10.76 | 2.57 | 12.1 | 1.84 | 0.04 | -0.07 | 0.12 | 0.73 | - | 1.09 | 0.30 | - | 4 | 0.05 | - |
| | T2 | 10.64 | 3.17 | 12.24 | 2.07 | | | | | | | | | | | |
| DKEFS - Condition 4 (scaled score) | T1 | 10.96 | 3.21 | 11.52 | 3.09 | -0.11 | -0.12 | 0.01 | 0.92 | - | 1.46 | 0.23 | - | 0.86 | 0.35 | - |
| | T2 | 11.32 | 3.12 | 11.9 | 2.79 | | | | | | | | | | | |
| *Verbal memory* | | | | | | | | | | | | | | | | |
| Free Recall (z-score) | T1 | -0.51 | 1.26 | -0.24 | 1.13 | -0.46 | -0.07 | 2.4 | 0.13 | - | 4.29 | 0.05 | - | 0.01 | 0.92 | - |
| | T2 | 0.03 | 0.96 | -0.17 | 1.11 | | | | | | | | | | | |
| Delayed Free Recall (z-score) | T1 | 0.05 | 1.45 | 0.15 | 1.47 | 0.03 | -0.22 | 1.43 | 0.23 | - | 0.89 | 0.35 | - | 0.37 | 0.54 | - |
| | T2 | 0 | 1.75 | 0.48 | 1.44 | | | | | | | | | | | |
| *Visual-spatial abilities* | | | | | | | | | | | | | | | | |
| MCTF - Copy (scaled score) | T1 | 31.69 | 3.92 | 31.17 | 3.68 | 0.18 | -0.62 | 6.02 | **0.01** | 0.16 | 1.08 | 0.30 | - | 0.72 | 0.39 | - |
| | T2 | 31.02 | 3.71 | 33.21 | 2.22 | | | | | | | | | | | |
| MCTF - Copy (z-score SES) | T1 | 0.11 | 0.98 | -0.06 | 1 | 0.17 | -0.62 | 5.86 | **0.02** | 0.16 | 1.49 | 0.22 | - | 0.71 | 0.40 | - |
| | T2 | -0.06 | 0.99 | 0.5 | 0.61 | | | | | | | | | | | |
| MCTF - Copy (z-score All) | T1 | -0.01 | 0.97 | -0.18 | 1 | 0.16 | -0.6 | 5.27 | **0.03** | 0.16 | 1.55 | 0.22 | - | 0.29 | 0.59 | - |
| | T2 | -0.16 | 0.98 | 0.37 | 0.64 | | | | | | | | | | | |
| MCTF - Recall (scaled score) | T1 | 15.1 | 5.12 | 16.52 | 5.84 | -0.1 | -0.2 | 0.28 | 0.60 | - | 1.42 | 0.24 | - | 1.44 | 0.24 | - |
| | T2 | 15.96 | 4.81 | 17.83 | 6.47 | | | | | | | | | | | |
| MCTF - Recall (z-score SES) | T1 | -0.02 | 0.91 | 0.17 | 1.08 | -0.11 | -0.2 | 0.27 | 0.61 | - | 1.49 | 0.23 | - | 0.98 | 0.33 | - |
| | T2 | 0.15 | 0.88 | 0.4 | 1.19 | | | | | | | | | | | |
| MCTF - Recall (z-score All) | T1 | -0.2 | 1.15 | -0.2 | 1.18 | -0.05 | 0.02 | 0.04 | 0.84 | - | 0.01 | 0.92 | - | 0.02 | 0.89 | - |
| | T2 | -0.12 | 1.12 | -0.22 | 1.21 | | | | | | | | | | | |

DSST, Digit Symbol Substitution Test; DKEFS, Delis-Kaplan executive function system (Stroop); FCSRT, French adaptation of the 16-items free and cued selective reminding test; MMSE, Mini Mental State Examination; MTCF/ROCF, Rey complex figure & modified Taylor complex figure tests; PPT, Purdue Pegboard Test; SES, sociodemographic variables (age, education level, and sex); TMT,Trail making test; WPo, sedative-hypnotics withdrawal plan group; WP+CBTi, CBTi combined to sedative-hypnotics withdrawal plan

Regarding cognitive evaluation, 25 participants (attrition rate 7.4%) from the WP+CBTi group and 21 participants (attrition rate 4.6%) from the WPo group completed overnights at 16 weeks post-randomization (T2).

#### Table S3: Spindle characteristics in central region

| | | WP+CBTi group | | WPo group | | Cohen g' (T1 vs T2) | | Time*Group | | | Time | | | Group | | |
|---|---|---|---|---|---|---|---|---|---|---|---|---|---|---|---|---|
| Outcome measure | (N) | Mean | SD | Mean | SD | WP+CBTi | WPo | F | *p* | *q* | F | *p* | *q* | F | *p* | *q* |
| **Spindles adapted characteristics** | | | | | | | | | | | | | | | | |
| (23 WP+CBTi vs 20 WPo) | | | | | | | | | | | | | | | | |
| ***NREM*** | | | | | | | | | | | | | | | | |
| Density | T1 | 1.19 | 0.24 | 1.15 | 0.18 | 0.21 | 0.41 | 0.02 | 0.88 | - | 5.97 | **0.02** | 0.06 | 0.1 | 0.74 | - |
| | T2 | 1.13 | 0.36 | 1.07 | 0.22 | | | | | | | | | | | |
| Duration | T1 | 0.76 | 0.05 | 0.76 | 0.04 | 0.48 | 0.72 | 0.36 | 0.54 | - | 9.72 | 0.002 | **0.04*** | 0.25 | 0.61 | - |
| | T2 | 0.74 | 0.03 | 0.73 | 0.03 | | | | | | | | | | | |
| Amplitude | T1 | 84.03 | 25.71 | 75.66 | 16.89 | 0.13 | -0.28 | 2.72 | 0.11 | - | 0.19 | 0.66 | - | 0.45 | 0.51 | - |
| | T2 | 80.89 | 21.81 | 81.09 | 20.07 | | | | | | | | | | | |
| Frequency | T1 | 11.4 | 1.15 | 11.3 | 1.19 | -0.25 | -0.27 | 0.03 | 0.86 | - | 2.09 | 0.16 | - | 0.03 | 0.87 | - |
| | T2 | 11.75 | 1.48 | 11.74 | 1.86 | | | | | | | | | | | |
| ***NREM2*** | | | | | | | | | | | | | | | | |
| Density | T1 | 1.13 | 0.25 | 1.09 | 0.19 | 0.2 | 0.3 | 0.36 | 0.55 | - | 4.25 | **0.04** | 0.13 | 0.1 | 0.76 | - |
| | T2 | 1.07 | 0.33 | 1.02 | 0.23 | | | | | | | | | | | |
| Duration | T1 | 0.77 | 0.06 | 0.76 | 0.03 | 0.47 | 0.63 | 0.07 | 0.79 | - | 6.18 | **0.01** | 0.06 | 0.27 | 0.60 | - |
| | T2 | 0.75 | 0.04 | 0.74 | 0.03 | | | | | | | | | | | |
| Amplitude | T1 | 79.69 | 24.92 | 72.38 | 15.11 | 0.1 | -0.32 | 2.48 | 0.12 | - | 0.47 | 0.49 | - | 0.25 | 0.62 | - |
| | T2 | 77.21 | 21.16 | 78.66 | 21.01 | | | | | | | | | | | |
| Frequency | T1 | 11.58 | 1.16 | 11.39 | 1.27 | -0.21 | -0.25 | 0.06 | 0.81 | - | 1.68 | 0.20 | - | 0.11 | 0.74 | - |
| | T2 | 11.87 | 1.52 | 11.82 | 1.89 | | | | | | | | | | | |
| ***NREM3*** | | | | | | | | | | | | | | | | |
| Density | T1 | 1.29 | 0.32 | 1.24 | 0.21 | 0.16 | 0.45 | 0.03 | 0.85 | - | 4.85 | **0.03** | 0.10 | 0.18 | 0.67 | - |
| | T2 | 1.23 | 0.43 | 1.13 | 0.29 | | | | | | | | | | | |
| Duration | T1 | 0.74 | 0.04 | 0.75 | 0.06 | 0.4 | 0.67 | 0.76 | 0.38 | - | 8.68 | 0.003 | **0.04*** | 1.14 | 0.29 | - |
| | T2 | 0.73 | 0.04 | 0.72 | 0.03 | | | | | | | | | | | |
| Amplitude | T1 | 89.08 | 28.92 | 79.51 | 19.33 | 0.07 | -0.13 | 0.6 | 0.44 | - | 0.02 | 0.90 | - | 1.07 | 0.31 | - |
| | T2 | 87.17 | 27.35 | 82.16 | 21.11 | | | | | | | | | | | |
| Frequency | T1 | 11.18 | 1.15 | 11.13 | 1.14 | -0.32 | -0.3 | 0.01 | 0.90 | - | 2.83 | 0.1 | - | <0.01 | 0.96 | - |
| | T2 | 11.6 | 1.4 | 11.62 | 1.85 | | | | | | | | | | | |

WPo, sedative-hypnotics withdrawal plan group; WP+CBTi, CBTi combined to sedative-hypnotics withdrawal plan

### Figures

#### Figure S1: Lower sedative-hypnotic dosage at baseline were associated with withdrawal success following intervention

*Sedative-hypnotic dose at baseline is negatively associated with the percentage reduction in self-reported consumption in the WP+CBTi group only.*

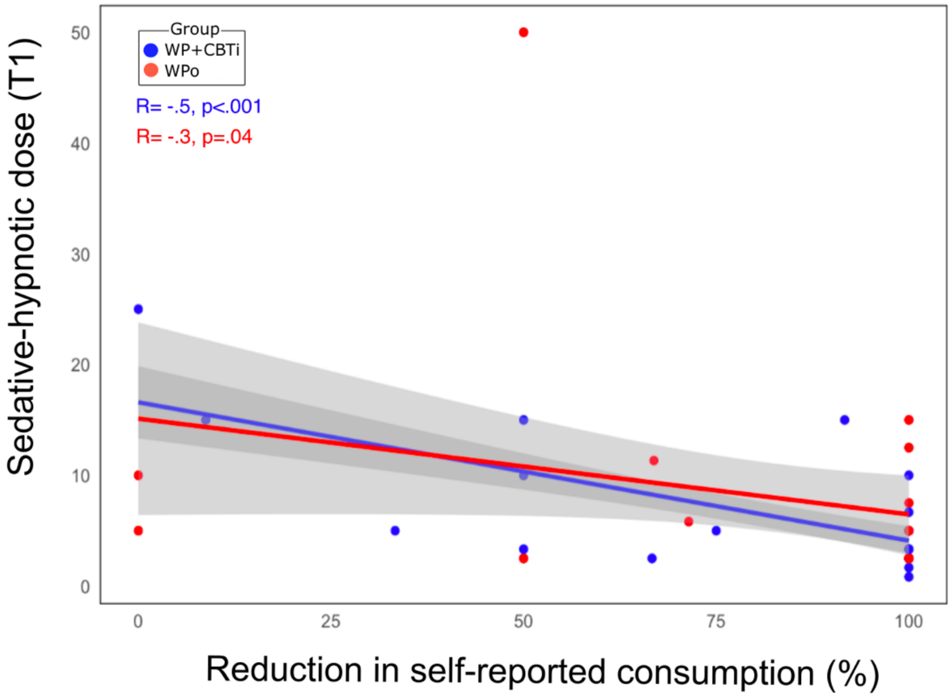

#### Figure S2: Association between spindle density reduction in central region and decreased sedative-hypnotic dose

*The decrease in central spindle density in WPo correlated with the reduction in the sedative-hypnotic dose consumed. However, similar correlation was not found in the WP+CBTi group.*

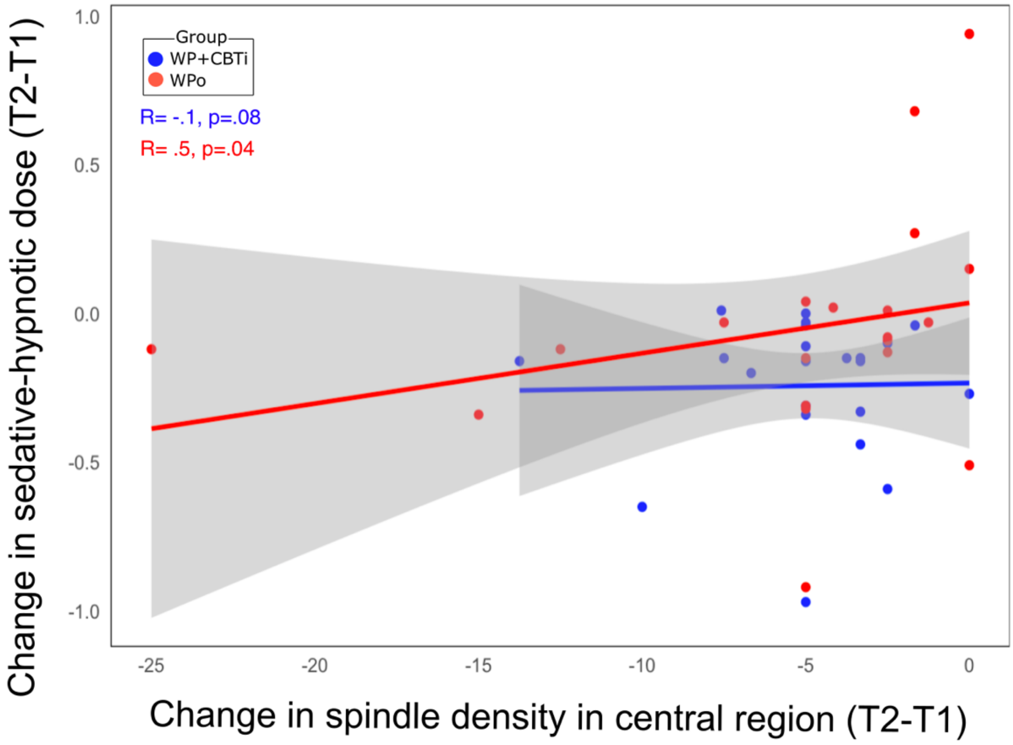
